# Impact of Individual and Contextual Factors on Measles, Mumps, and Rubella Vaccination in Brazil: A Multilevel Analysis

**DOI:** 10.1101/2025.09.30.25337024

**Authors:** Tatiana Lang D’Agostini, Fernanda Florencia Fregnan Zambom, Ana Paula França, Manoel Carlos Sampaio de Almeida Ribeiro, Rita Barradas Barata

**Affiliations:** Department of Public Health, Santa Casa de São Paulo Faculty of Medical Sciences, São Paulo, SP, Brazil; “Prof. Alexandre Vranjac” Center of Epidemiological Surveillance, Disease Control Coordination, São Paulo, SP, Brazil

**Keywords:** Vaccination coverage, MMR vaccine, Social determinants of health, Multilevel analysis, Child immunization

## Abstract

Vaccination is a crucial public health intervention, and Brazil’s National Immunization Program (PNI) has historically achieved high coverage rates. However, since 2014, and especially during the COVID-19 pandemic, MMR (measles, mumps, and rubella) vaccine coverage has declined significantly. In this context, understanding how individual and contextual factors contribute to incomplete vaccination is vital to reversing this trend and preventing disease resurgence. The objective of this study was to analyze the influence of individual, household, and contextual factors on incomplete MMR vaccination in Brazilian children, using a multilevel analytical approach. This study analyzed secondary data from the 2020 National Immunization Coverage Survey, which assessed MMR vaccination among children born in 2017 and 2018. The survey included 26 state capitals, the Federal District, and 12 mid-sized municipalities. Multilevel logistic regression models were used to identify factors associated with the absence of a valid second MMR dose. The absence of a second MMR dose was significantly associated with higher birth order, maternal employment, and not having a partner. Maternal education showed a protective effect, especially among mothers with higher education levels. Participation in the Bolsa Família program reduced the likelihood of incomplete vaccination. Unexpectedly, children in lower socioeconomic strata (C and D) had lower odds of incomplete vaccination than those in wealthier strata, suggesting effective targeting of public policies. Most of the variance occurred at the Primary Sampling Unit level, highlighting the importance of local health infrastructure. Incomplete MMR vaccination in Brazil is influenced by both individual and contextual factors. Equity-focused strategies that address maternal workload, family structure, and local service delivery are essential to improving coverage and preventing the resurgence of vaccine-preventable diseases.

## Introduction

Vaccination is considered by the World Health Organization (WHO) to be one of the most effective public health strategies, with a significant impact on the prevention of infectious diseases [1]. In Brazil, the National Immunization Program (PNI), established in 1973, has become a global reference, coordinating immunization efforts across the national territory [2].

The introduction of the measles vaccine took place in the 1960s, but its impact was significantly enhanced with the implementation of the PNI [3,4]. The State of São Paulo (SSP) stood out in 1987 by launching the first mass measles vaccination campaign, inspired by the Cuban experience of measles vaccination [5,6]. The campaign achieved 91% coverage among children aged 9 months to 14 years, leading to a 98% reduction in disease incidence and the elimination of measles-related deaths. In 1992, Brazil launched the National Measles Elimination Plan, conducting high-coverage campaigns that reached 96% of the target population [7].

According to the Brazilian immunization schedule, measles vaccination requires two doses administered between 12 and 15 months of age. However, since 2014, Brazil has failed to meet the 95% coverage target recommended by WHO [1]. Despite efforts involving the MMR vaccine (measles, mumps, and rubella), outbreaks have reemerged. In 1996, the SSP experienced an epidemic with over 42,000 cases and 42 deaths. Although endemic transmission was interrupted in 2000, measles was reintroduced in 2018 following an outbreak that began in Venezuela [8,9].

Regarding rubella, outbreaks between 2006 and 2008 resulted in over 6,700 cases, prompting a national vaccination campaign in 2008 that achieved 97% coverage. Transmission was interrupted in 2009, and Brazil was declared free of rubella and Congenital Rubella Syndrome (CRS) in 2015. However, declining vaccination coverage now threatens these achievements. Barriers such as limited access to remote regions, failures in information systems, and vaccine hesitancy, fueled by misinformation, jeopardize the sustained elimination of these diseases [3, 10].

Since 2016, Brazil has experienced a consistent decline in vaccination coverage, particularly for BCG, poliomyelitis, and MMR. The COVID-19 pandemic in 2020 and 2021 exacerbated this trend, leading to a drop of up to 65% in childhood vaccinations. Data from the Ministry of Health show that while vaccination coverage exceeded 100% in 2013 and 2014, it had fallen to 91.2% by 2019 [2, 9, 11-13].

Globally, although MMR vaccine coverage remains above 80% for the first dose (D1) and 70% for the second dose (D2), there was a rise in measles cases in 2022. In the Americas, data from 2024 and early 2025 indicate a significant increase in cases, particularly in the United States and Canada. In Brazil, although the last endemic case of measles was reported in 2022, the country remains classified as “pending re-verification” for elimination status [14,15].

Vaccination coverage is the result of a complex interaction between individual, familial, and contextual factors. The adherence to childhood immunization schedules is strongly influenced by maternal characteristics, such as education level, employment status, and household structure. Studies have shown that lower maternal education is associated with reduced likelihood of completing the child’s vaccination series, while higher birth order correlates with increased risk of incomplete immunization, likely due to limited parental time, resources, and attention [16-18].

Socioeconomic disparities further exacerbate vaccination gaps by limiting access to healthcare services and influencing health-related decision-making. Families in situations of poverty often face logistical challenges, such as transportation barriers, inflexible work schedules, and insufficient community support, which may hinder timely vaccine administration [19,20].

Moreover, contextual factors, including the organization of primary care services, quality of local health infrastructure, and communication strategies, are critical in shaping vaccination outcomes. In recent years, vaccine hesitancy has emerged as a growing public health concern, driven by misinformation, declining risk perception, and distrust in government institutions. This phenomenon has been particularly evident in urban and wealthier populations, posing new challenges to the effectiveness of immunization programs [21-24].

These multifaceted barriers underscore the necessity of equity-focused policies and intersectoral strategies that address both demand- and supply-side determinants of vaccination. Considering Brazil’s recent declines in immunization coverage, particularly for the MMR vaccine, it is imperative to understand how individual and structural factors influence immunization behavior to inform more targeted and effective interventions.

## Materials and Methods

This is a study based on secondary data obtained from a population-based household survey designed to assess MMR vaccine coverage. The study is part of the 2020 National Immunization Coverage Survey [25], conducted using a probabilistic sample representative of the cohort of live births from 2017 to 2018. The sample included the 26 state capitals, the Federal District, and 12 mid-sized municipalities (with over 100,000 inhabitants) located outside metropolitan regions. The 2020 National Immunization Coverage Survey [25] data were accessed between September 2020 and March 2022, ensuring that all children included were at least 24 months of age at the time of the survey. The estimate of MMR vaccination coverage, delays, and loss to follow-up among children up to 24 months old is reported in D’Agostini et al. [26].

For statistical analysis, multilevel logistic regression modeling was employed using the IBM SPSS Modeler 15.0 Software, considering the following hierarchical levels:

### 1. Municipal Level

Comprised of 39 geographic units, including the state capitals and the Federal District.

### 2. Socioeconomic Stratum Level

Each municipality was subdivided into four homogeneous socioeconomic strata (A, B, C, and D), defined based on aggregated indicators and formed by clusters of Primary Sampling Units (PSUs).

### 3. Primary Sampling Unit (PSU) Level

PSUs consisted of one or more census tracts randomly selected within each stratum.

### 4. Individual Level

Within each PSU, individuals were randomly selected to comprise the analysis sample. The individual level represents the final observational unit of the study.

The dependent variable was defined as the absence of a valid second dose of the MMR vaccine (binary outcome). First-level independent variables included demographic, socioeconomic, and household characteristics of individuals.

The statistical model was structured to include random effects associated with PSU, socioeconomic stratum, and municipality levels, capturing residual variability at each of these levels. Three models were progressively fitted, with the full model represented by the following equation:

logit(P(Yij=1))=β0+β1X1+β2X2+…+μupa[j]+μstratum[k]+μmunicipality[l]+ϵij\text{ logit}(P(Y_{ij} = 1)) = \beta_0 + \beta_1X_1 + \beta_2X_2 + \ldots +

\mu_{\text{upa}[j]} + \mu_{\text{stratum}[k]} + \mu_{\text{municipality}[l]} +

\epsilon_{ij}logit(P(Yij=1))=β0+β1X1+β2X2+…+μupa[j]+μstratum[k]

+μmunicipality[l]+ϵij

Where:

- YijY_{ij}Yij: response variable (non-receipt of the second dose);
- β0\beta_0β0: intercept;
- β1X1+β2X2+…\beta_1X_1 + \beta_2X_2 + \ldotsβ1X1+β2X2+…: individual-level covariates;
- μupa[j],μstratum[k],μmunicipality[l]\mu_{\text{upa}[j]}, \mu_{\text{stratum}[k]}, \mu_{\text{municipality}[l]}μupa[j],μstratum[k],μmunicipality[l]: random variance components for PSU, stratum, and municipality, respectively;
- ϵij\epsilon_{ij}ϵij: individual-level random error.

## Ethics Statement

This study use secudary data from the National Vaccine Coverage Survey 2020 [25], approved by the Research Ethics Committee of the Institute of Collective Health at the Federal University of Bahia (approval no. 3.366.818, issued on June 4, 2019; Certificate of Presentation for Ethical Consideration [CAAE] 4306919.5.0000.5030) and by the Research Ethics Committee of the Brotherhood of Santa Casa de São Paulo (approval no. 4.380.019, issued on November 4, 2020; CAAE 39412020.0.0000.5479), as well as by the Scientific Committee of the Department of Public Health at the Faculty of Medical Sciences, Santa Casa de São Paulo (opinion no. 03/2024).

The study posed minimal risk to the target population during or after data collection. This study used secondary data obtained from the National Vaccine Coverage Survey 2020. All analyses were conducted in compliance with the Brazilian General Data Protection Law (Law No. 13.709/2018). The dataset does not contain nominal information, thus preventing the identification of individual participants, and all data were handled under strict confidentiality.

## Results

In the multilevel analysis of incomplete MMR vaccination, the null model was first estimated to assess the variance in the outcome (defined as the absence of two valid doses of the MMR vaccine) across hierarchical levels, without including covariates. Subsequently, Model 2 incorporated individual-level variables and adjusted the random effects to better explain the observed variability. Finally, Model 3 added intramunicipal socioeconomic strata (categories A, B, C, and D) as fixed effects, while maintaining the previously tested individual-level predictors (Table 1).

**Table 1.** Random Effect, Intraclass Correlation Coefficients and Model Fit Statistics, Brazil, 2025.

|  | Null Model | Model 2 | Model 3 |
| --- | --- | --- | --- |
| **Random Effects** |  |  |  |
| Variance – Municipality | 0.1039 | 0.1056 | 0.1094 |
| Variance – Stratum | 0.0650 | 0.0766 | 0.0557 |
| Variance – PSU | 0.3389 | 0.3386 | 0.3424 |
| **Intraclass Correlation Coefficients (ICC)** |  |  |  |
| Municipality | 2.74% | 2.77% | 2.88% |
| Stratum | 4.45% | 4.78% | 4.35% |
| PSU | 13.37% | 13.67% | 13.36% |
| **Model Fit Statistics** |  |  |  |
| Log-Likelihood | -17817.18 | -16701.55 | -16691.99 |
| AIC | 35642.36 | 33435.10 | 33421.98 |
| BIC | 35676.52 | 33571.02 | 33583.38 |

The analysis of random effects revealed a slight increase in the variance between municipalities in Models 2 and 3, suggesting that the inclusion of covariates made contextual differences between cities more evident. Regarding socioeconomic strata, variance increased in Model 2 and decreased in Model 3 after the stratum was included as a fixed effect. This pattern indicates that part of the variability across strata can be explained by the added variables. The variance at the Primary Sampling Unit (PSU) level remained stable across all models, indicating that this level represents the main source of variation in the outcome (Table 1).

The Intraclass Correlation Coefficient (ICC) estimates reinforced these findings. The ICC for the municipal level showed a slight increase in the subsequent models, highlighting the growing influence of local contextual characteristics as other sources of variability are controlled. The ICC for the socioeconomic strata increased in Model 2 and decreased in Model 3, supporting the conclusion that the inclusion of the stratum as a fixed effect accounts for a significant portion of the heterogeneity across socioeconomic groups. The ICC for the PSUs remained nearly constant, underscoring the importance of this level as a determinant of differences in vaccination patterns.

Model fit indicators, such as the log-likelihood, improved progressively, indicating a better fit of the models to the data with the addition of predictors. Similarly, information criteria (AIC and BIC) decreased across models, confirming increased parsimony and precision in the more refined models (Table 1).

Among the individual-level predictors, the following were significantly associated with incomplete vaccination: birth order, presence of a partner, maternal employment status, and participation in the Bolsa Família Program (a direct cash transfer program with conditionalities that benefits families living in poverty and extreme poverty). These factors consistently showed significant associations across all models, suggesting a robust influence on the outcome. Maternal education remained a protective factor, although its effect was slightly attenuated in Model 3, likely due to overlap with the socioeconomic stratum variable. Likewise, the influence of consumption level was partially explained by the inclusion of the stratum, emphasizing the role of socioeconomic inequalities in shaping vaccination coverage patterns (Table 2).

**Table 2.** Multilevel Logistic Regression Models for Incomplete MMR Vaccination, Brazil, 2025.

| Description/ Fixed Effects | Null Model | Model 2 | Model 3 |
| --- | --- | --- | --- |
| **Birth Order** | - |  |  |
| First | - | 1 (ref) | 1 (ref) |
| Second | - | 1.16 (1.09–1.24) | 1.17 (1.09–1.24) |
| Third | - | 1.45 (1.32–1.58) | 1.45 (1.32–1.58) |
| Fourth or more | - | 1.62 (1.44–1.82) | 1.62 (1.44–1.82) |
| **Mother has a partner** | - |  |  |
| Yes | - | 1 (ref) | 1 (ref) |
| No | - | 1.16 (1.08–1.24) | 1.16 (1.08–1.24) |
| **Mother is employed** | - |  |  |
| Yes | - | 1.16 (1.09–1.23) | 1.16 (1.09–1.23) |
| No | - | 1 (ref) | 1 (ref) |
| **Mother's education** | - |  |  |
| Up to 8 years | - | Reference | Reference |
| 9 to 12 years | - | 1.04 (0.93–1.18) | 1.04 (0.92–1.17) |
| 13 to 15 years | - | 0.91 (0.81–1.02) | 0.91 (0.81–1.02) |
| 16 or more years | - | 0.84 (0.73–0.97) | 0.83 (0.72–0.96) |
| **Consumption Level** | - |  |  |
| A (≥ 42 points) | - | 1 (ref) | 1 (ref) |
| B (27–41 points) | - | 0.81 (0.71–0.94) | 0.83 (0.72–0.96) |
| C (16–26 points) | - | 0.82 (0.70–0.96) | 0.86 (0.74–1.00) |
| D (≤ 15 points) | - | 0.88 (0.76–1.05) | 0.93 (0.78–1.10) |
| **Bolsa Família Program** | - |  |  |
| Yes | - | 0.81 (0.75–0.88) | 0.81 (0.75–0.88) |
| No | - | 1 (ref) | 1 (ref) |
| **Stratum** | - | Not included |  |
| A | - | - | 1 (ref) |
| B | - | - | 0.88 (0.76–1.02) |
| C | - | - | 0.74 (0.64–0.86) |
| D | - | - | 0.74 (0.64–0.87) |

ref, reference.

Finally, the inclusion of the socioeconomic stratum as a fixed effect in Model 3 revealed that strata C and D had a lower probability of incomplete vaccination compared to stratum A.

## Discussion

The present study offers a comprehensive multilevel analysis of incomplete MMR vaccination coverage in Brazil, contributing to the growing body of literature on the social determinants of immunization. By integrating individual, household, and contextual factors, this analysis provides important insights into the mechanisms that underlie vaccination disparities and highlights priority areas for public health interventions.

Our findings align with and extend results from both Brazilian and international studies, particularly in demonstrating how family structure, maternal characteristics, and socioeconomic context influence incomplete MMR vaccination. We observed graduated increases in odds of incomplete vaccination, from second-born (aOR ≈ 1.17) to fourth-born or later (aOR ≈ 1.62). Similar patterns were identified in a U.S. national study, where non-first-born children were less likely to be fully up-to-date, even after adjusting for family size, with authors suggesting parental time constraints and lowered urgency for subsequent children as possible explanations [16]. A global survey across 85 low- and middle-income countries similarly showed zero-dose prevalence rising from 11.0% in firstborns to 17.1% in fifth-born or higher [27].

Higher maternal education proved protective, particularly in the highest education categories (16+ years: OR ≈ 0.83). This accords with a 2017 meta-analysis showing children of mothers with secondary or higher education had 2.3 times greater odds of completing vaccination [17]. Another systematic review in Ethiopia found maternal primary and secondary education essentially doubled or tripled vaccination completion rates [18]. However, in our Model 3, the effect of maternal education was attenuated by the inclusion of socioeconomic stratum, suggesting part of its influence operates through broader socioeconomic conditions.

Interestingly, children in lower socioeconomic strata (C and D) were less likely to have incomplete vaccinations compared to those in the wealthiest stratum A. This mirrors earlier Brazilian research, notably a 2012 survey across 27 capitals, which found children in wealthier census tracts exhibited lower complete vaccination coverage [19]. In Londrina (Paraná), coverage was lowest in stratum A (36%) and highest in stratum D (70%) [28], suggesting more active public health interventions or perceptions of risk influence uptake in lower-income groups.

Our finding that participation in Bolsa Família reduced the likelihood of incomplete vaccination (OR ≈ 0.81) supports the program’s documented role in decreasing health inequities. Moreover, maternal employment and absence of a partner increased incompleteness, indicating constraints related to time, childcare responsibilities, and healthcare access, findings consistent with broader literature on social determinants of immunization [19].

Additional factors such as household income, maternal age, access to transportation, availability of flexible clinic hours, and previous negative experiences with healthcare services have also been linked to lower vaccination rates [29-32]. Cultural beliefs and lack of knowledge about vaccination schedules can further contribute to delays or missed doses, particularly in rural or underserved communities [33].

The PSU level contributed most of the outcome variability (ICC ∼13%), with municipal and stratum-level ICCs remaining modest (<5%). This pattern highlights heterogeneity within health service delivery units, likely administrative or geographical areas, aligning with studies that emphasize local health infrastructure as a strong determinant of vaccination [34].

Recent disruptions caused by the COVID-19 pandemic have further exacerbated vaccination gaps globally and in Brazil, particularly among vulnerable populations, with childhood immunization rates declining sharply in many regions [23, 24, 35]. In addition, vaccine hesitancy, driven by misinformation and declining risk perception, has emerged as a growing challenge to immunization programs [21-22].

In this context, studies emphasize the importance of community-based strategies and intersectoral collaboration to improve vaccine uptake, especially in hard-to-reach populations [20, 36, 37]. These findings reinforce the need for targeted, equity-oriented policies to rebuild confidence in vaccines and mitigate structural barriers to access.

## Conclusion

This study highlights that individual and contextual factors influence incomplete MMR vaccine coverage in Brazil. The use of a multilevel approach revealed that higher birth order, maternal employment, and the absence of a partner increase the chances of incomplete vaccination, while higher maternal education and participation in the Bolsa Família program protect against this risk. The significant variation between census units reinforces the importance of local context and the organization of health services.

The lower incidence of incomplete vaccination in lower-income families indicates that targeted public policies may be working. Therefore, it is urgent to implement intersectoral, equity-focused strategies to overcome barriers related to maternal workload, family structure, and access to health care, ensuring broad vaccination coverage and preventing the return of preventable diseases.

## Data Availability

The raw data cannot be publicly shared due to ethical restrictions and the inclusion of sensitive personal information. However, researchers may request access to the raw data from the 2020 National Immunization Coverage Survey. Data access requests should be directed to José Cássio de Moraes, Principal Investigator of the project, by e-mail at.

## Acknowledgment

To the 2020 National Vaccination Coverage Survey group, especially to the principal investigator, Professor José Cássio de Moraes.

## Notes

### Competing Interest Statement

The authors have declared no competing interest.

### Funding Statement

The author(s) received no specific funding for this work.

### Author Declarations

The 2020 National Immunization Coverage Survey was approved by the Research Ethics Committees involving Human Subjects of the Institute of Collective Health at the Federal University of Bahia (approval nº 3.366.818, June 4, 2019 Certificate of Ethical Appreciation Submission – CAAE 4306919.5.0000.5030) and of the Irmandade da Santa Casa de São Paulo (approval nº 4.380.019, November 4, 2020 CAAE 39412020.0.0000.5479). This study was also approved by the Scientific Committee of the Department of Public Health, Faculty of Medical Sciences, Santa Casa de São Paulo (approval nº 03/2024).

